# Effectiveness of portable air filtration on reducing indoor aerosol counts: preclinical observational trials

**DOI:** 10.1101/2021.04.26.21256152

**Authors:** Jung Hoon Lee, Max Rounds, Forbes McGain, Robyn Schofield, Grant Skidmore, Imogen Wadlow, Kevin Kevin, Ashley Stevens, Caroline Marshall, Lou Irving, Marion Kainer, Kirsty Buising, Jason Monty

## Abstract

**Objective:** To assess the effectiveness of aerosol filtration by portable air cleaning devices with high efficiency particulate air (HEPA) filters used in addition to standard building heating ventilation and air-conditioning (HVAC).

**Methods:** Test rooms, including a hospital single-patient room, were filled with test aerosol to simulate aerosol movement. Aerosol counts were measured over time with various portable air cleaning devices and room ventilation systems to quantify the aerosol concentration reduction rate and overall clearance rate.

**Results:** Portable air cleaners were very effective in removing aerosols, especially for the devices with high flow rate. In a small control room, the aerosols were cleared 4 to 5 times faster with portable air cleaners than the room with HVAC alone. A single bed hospital room equipped with an excellent ventilation rate (∼ 14 air changes per hour) can clear the aerosols in 20 minutes. However, with the addition of two air cleaners, the clearance time became 3 times faster (in 6 minutes and 30 seconds).

**Conclusions:** Portable air cleaning devices with HEPA filtration were highly effective at removing aerosols. To clear aerosols (above 90% clearance) in under 10 minutes requires around 25 air changes per hour; readily feasible with air cleaners. Inexpensive portable air cleaning devices should be considered for small and enclosed spaces in health care settings such as inpatient rooms, personal protective equipment donning/doffing stations, and staff tea rooms. Portable air cleaners are particularly important where there is limited ability to reduce aerosol transmission with building HVAC ventilation.

## Introduction

The range of possible transmission pathways of SARS-CoV2 and their relative contribution in various settings is still being investigated^1,2^, although the evidence for aerosol transmission seems robust.^3^ There is an emerging view that transmission via inhalation of aerosol particles probably plays a dominant role especially in indoor environments where the ventilation is poor and have airflow pathways that direct virus-laden air towards people.^4-7^ Engineering controls are needed to mitigate aerosol transmission in high risk settings including hospital wards, classrooms, and offices.

## Background

Building Heating, Ventilation, and Air Conditioning (HVAC) systems provide fresh/filtered air to a room at a controlled temperature for human comfort. At standard rates in hospital wards, the air change rates provided by the HVAC do little to mitigate the risk of aerosol transmission.^8^ Numerous existing policies and guidelines already suggest the use of portable HEPA filters to improve indoor air quality^9-11^, but there has been notably low uptake in the community.

Portable air cleaners with HEPA filtration clean the air of aerosol inside a room close to the infected person, providing *source control* and improve aerosol clearance rate beyond the HVAC system. It must be noted that not all “air change” methods are equal from an infection control viewpoint, especially for control of the aerosol transmission. The air exchange rate of HVAC is defined as the number of times that the volume of air in a room is replaced with fresh/filtered air introduced by the HVAC system in a given period. The commonly used measure of Air Changes per Hour of HVAC (ACH_HVAC_) is the ratio between volume flow rate of the system (*Q*_HVAC_) and volume of the room (*V*).

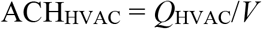

Portable air cleaning devices are designed to filter out aerosols (mainly intended for dust particles and other pollutants) from the air inside a room. Hence, the way that these devices provide “air changes” and reduce aerosol counts in a room is different to the HVAC system. i.e., HVAC pushes aerosol out of a room, while air cleaners filter out the aerosol from within the room. The air filtration rate per hour of portable air cleaning devices can be considered as equivalent air changes per hour, ACHe (Allen and Ibrahim 2021), and defined in a similar way to ACH_HVAC_, that is:

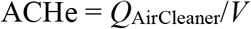

Where *Q*_AirCleaner_ is volume flow rate of air cleaning devices (also known as clean air delivery rate, CADR). Both of these “air change” methods reduce aerosol in indoor environments such that, in practice, it is possible to combine the two, such that air ventilation (ACH_HVAC_) and air filtration (ACHe) rates can be added into a total Air Changes per Hour (ACH) parameter for the purpose of infection control associated with aerosol transmission.^12,13^ Thus,

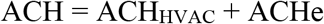

In this study, the single parameter ACH will be used to indicate the overall air change rate for reducing aerosol counts.

For practical purposes, predicting the clearance time for sufficiently high aerosol removal efficiency (e.g. above 90% clearance) is important to control the risk of aerosol transmission. An equation to estimate a clearance time for a given clearance rate with increasing ACH (ACH_HVAC_ + AFH) for aerosol removal is given by

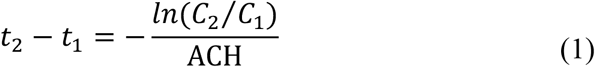

Where *t*_*1*_ and *t*_*2*_ are initial and final time stamps, *C*_*1*_ and *C*_*2*_ are initial and final concentration of aerosol. *t*_*2*_ − *t*_*1*_ is the clearance time with an assumption that the aerosols are homogenously mixed.^14^ This clearance time can be used to investigate the effectiveness of air ventilation and filtration systems for a given ACH.

The aim of this work is to assess the effectiveness of several common portable air cleaners to reduce aerosol particle counts, with simulated or actual building ventilation systems to demonstrate practical performance.

## Method

A full sized “control room” designed to duplicate the size of a standard single bedroom or office was constructed in the Michell Hydrodynamics Laboratory at the University of Melbourne. The room was well sealed and blackened for experimental flow visualisation purposes. The floor area of the room was 10.5m^2^ and the volume was approximately 24 m^3^. A schematic of the room is shown in figure 1(a). A fan with a flow rate of 55 m^3^/hr was used to deliver clean air (HEPA-filtered) through a ceiling duct with a conical diffuser installed to simulate standard HVAC (hereafter referred to as “control room HVAC”). A single exhaust vent for the air to leave the room was installed close to the ceiling replicating the common practice in indoor spaces. This airflow rate was calculated to be equivalent to 2.3 ACH for the tested control room. For experiments with an air cleaner, the portable unit was placed at one end of the room as shown in figure 1. A particle illumination technique with laser light was used to quantify the aerosol density in the room. The main purpose of this method is to acquire images of aerosol particles illuminated by the laser sheet. Since each particle reflects laser light to a camera, we can effectively (indirectly) count the number of particles illuminated by measuring the amount of light reflected by the particles within an acquired image. That is, a brighter image indicates more particles are in the plane of the laser sheet and a darker image indicates less particles. A laser sheet was created using a 150mW, 532 nm (green) laser and a cylindrical lens. A Canon XA40 digital camera was used as an imaging device. This particle illumination method is identical to the particle image velocimetry (PIV) technique which is widely used in experimental fluid mechanics^15^ and similar to the imaging system used in Bluyssen *et al*. (2021)^16^ to track aerosol counts. With no added aerosol to the control room, the aerosol particles were not visible through the camera suggesting the room was sufficiently clean of aerosol/dust particles prior to experimentation. Also, the preliminary measurement has shown (not included here for brevity) that only the reflected light from particles within the laser sheet affect the brightness of the image (i.e. negligible light is absorbed by particles between the laser sheet and the camera). For all experiments, theatrical smoke (aqueous glycol solution; the mean aerosol size of 1 µm) was injected for 15 seconds to introduce the aerosol particles into the room. The smoke particles were used as tracer aerosols to investigate aerosol movement within the room. Once the smoke particles were injected, an additional 10-minute waiting period was given for the smoke to mix and for transient flows due to injection to stabilize before each device was switched on and the experiments began. When aerosols were disseminated through the room the images showed the green laser sheet lighting up the smoke particles. Over time, as the air cleaners or HVAC cleared the smoke particles, the acquired images became less bright. The measured light intensity over a sub-region of the images as a function of time was acquired and the particle images were acquired until the illuminated aerosol was no longer visible by the camera.

**Figure 1:**
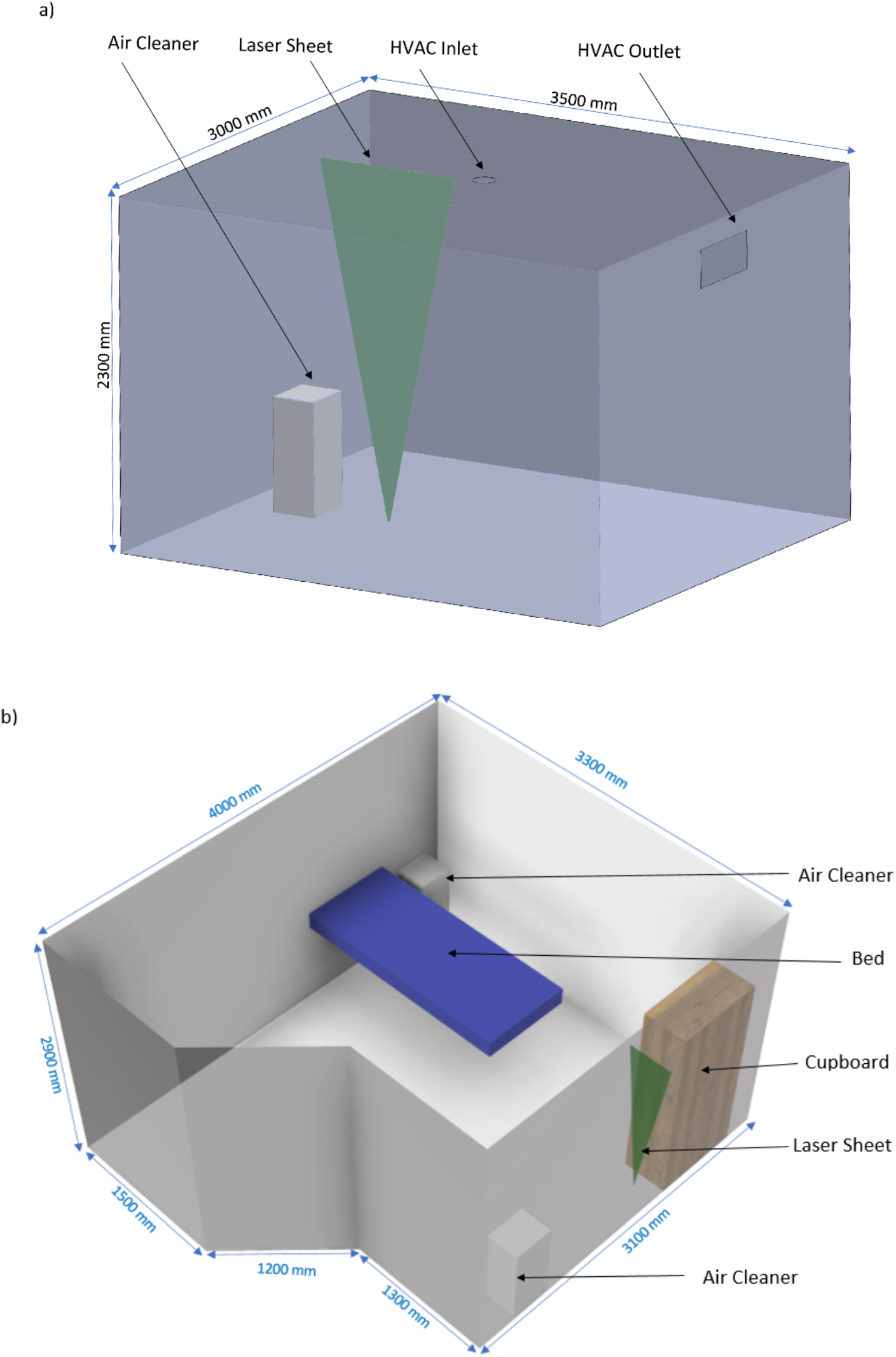
(a) Control room at The University of Melbourne and (b) Room geometry and illustration of laser-based smoke measurement setup in a single bed hospital room.

The same method was then used in a single bed patient room at the Royal Melbourne Hospital, shown in figure 1(b). The room floor space and volume were approximately 12 m^2^ and 37 m^3^ respectively and this room had HVAC with 13.9 ACH (hereafter referred to as “hospital HVAC”). This dataset was collected as part of a collaborative measurement campaign performed at the Royal Melbourne Hospital. Further details of the measurement campaign are available in Buising *et al*. (2021)^17^.

Three commercial air cleaners with three different inlet flow rates were tested in the control clean room in the laboratory. The flow rates (i.e. CADR) were approximately 200m^3^/hr (air cleaner A, Model: Industrial air cleaner 1, Westaflex, Australia), 400m^3^/hr (air cleaner B, Model: Industrial air cleaner 2, Westaflex, Australia) and 467 m^3^/hr (air cleaner C, Model: Air Purifier AX60RR5080WD, Samsung Electronics, South Korea). All tested air cleaners were equipped with H13 HEPA filters capable of filtering 99.97% of particles greater than 0.3µm.

Based on laboratory testing, for the size of the hospital room, two air cleaners (air cleaner C) were required to achieve a comparable air filtration rate to the control room measurement with one air cleaner C. The air cleaners were placed in regions that were close to a hospital bed and suspected of having poor air circulation by inspection. Contaminated particles could potentially stay in these regions longer than regions of the room with high circulation, increasing the risk of infection.

## Results

The configurations of the rooms tested are provided in table 1

**Table 1:**
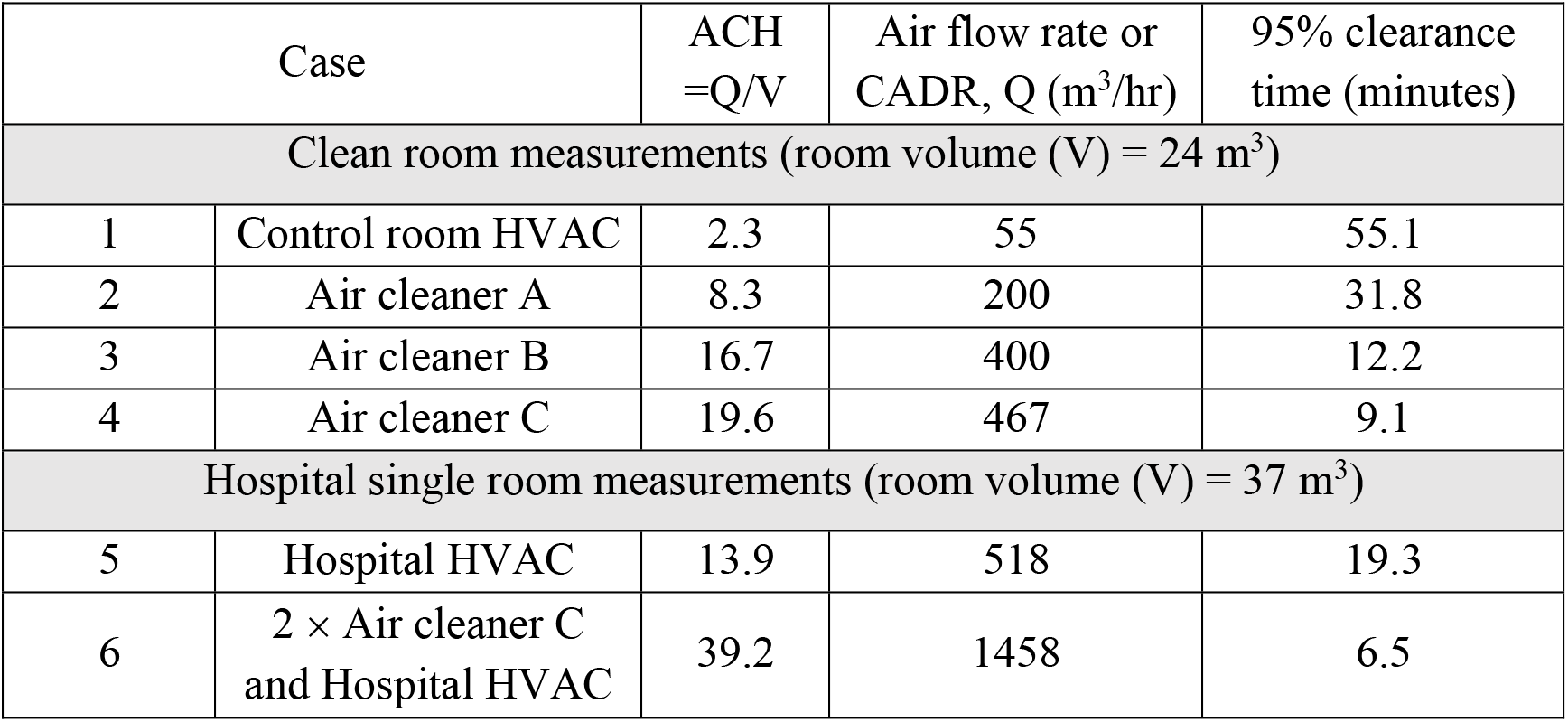
Summary of experimental configurations and parameters including tested room volumes, air flow rates and ACH.

Figure 2 shows the decrease of the aerosol particle density, *C*_smoke_(*t*) as a function of time for a range of ACH. Taking the initial light intensity as a measure of the initial density of the smoke particles, *C*_smoke_(0), the intensity data in figure 2 is computed by dividing the average light intensity associated with the number of illuminated particles at any given time by the initial intensity. Figure 2 shows a very clear decrease in aerosol clearance time with increasing ACH. This is important since it shows the effect of the air cleaner devices is to increase aerosol clearance rate at a given ACH.

**Figure 2:**
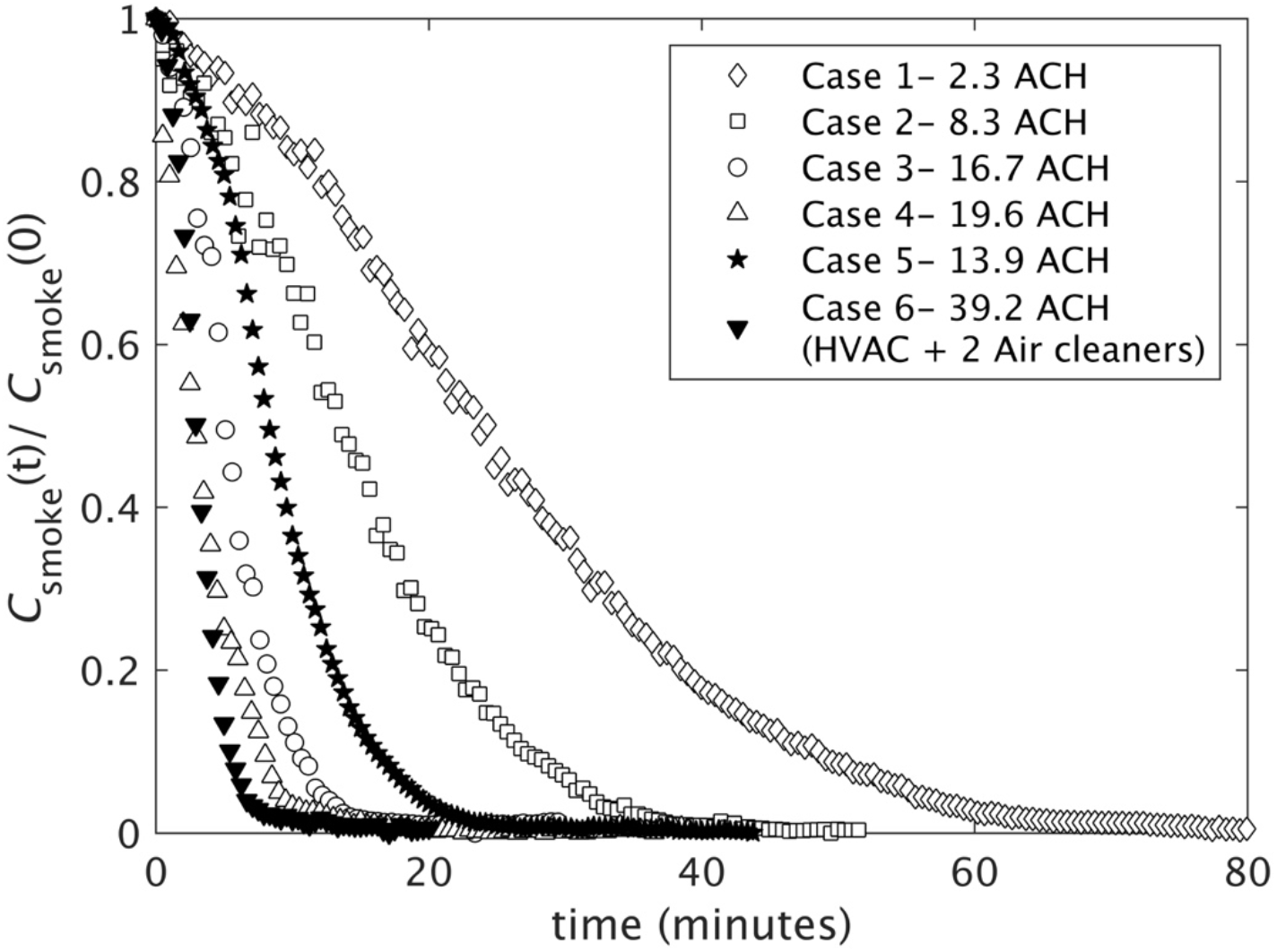
Smoke particle density decay with time. Aerosol particle density, *C*_smoke_(*t*), is measured as light intensity of images acquired and normalisation is by the initial light intensity, *C*_smoke_(0), which is smoke particle density. The white and black symbols indicate the smoke concentration decay rate measured in control room and real hospital room, respectively.

To further assess the practical performance of the portable air cleaners in reducing aerosols, the aerosol clearance time as a function of ACH is employed. Figure 3 shows the experimental results (symbols) from both the control and the hospital rooms for 63%, 90% and 95% clearances. The clearance time estimation profiles (lines) obtained from equation (1) at matched clearance rates to the experiments are plotted for comparison. The resulting clearance time from the experimental data clearly indicate that portable air cleaners with high flow rates (16.7 and 19.6 ACH) reduced the clearance time significantly. In the small control room, the aerosols were almost completely cleared 4 to 5 times faster (under 12 minutes) with portable air cleaners than the control room with HVAC alone (2.3 ACH). The low flow rate portable air cleaner with 8.3 ACH (case 3) was still significantly better than the HVAC. The hospital room with HVAC alone had a relatively high flow rate at baseline (13.9 ACH), however, when there were two air cleaners in the room (39.2 ACH in total) the clearance time was significantly improved to become 3 times faster (under 10 minutes).

**Figure 3:**
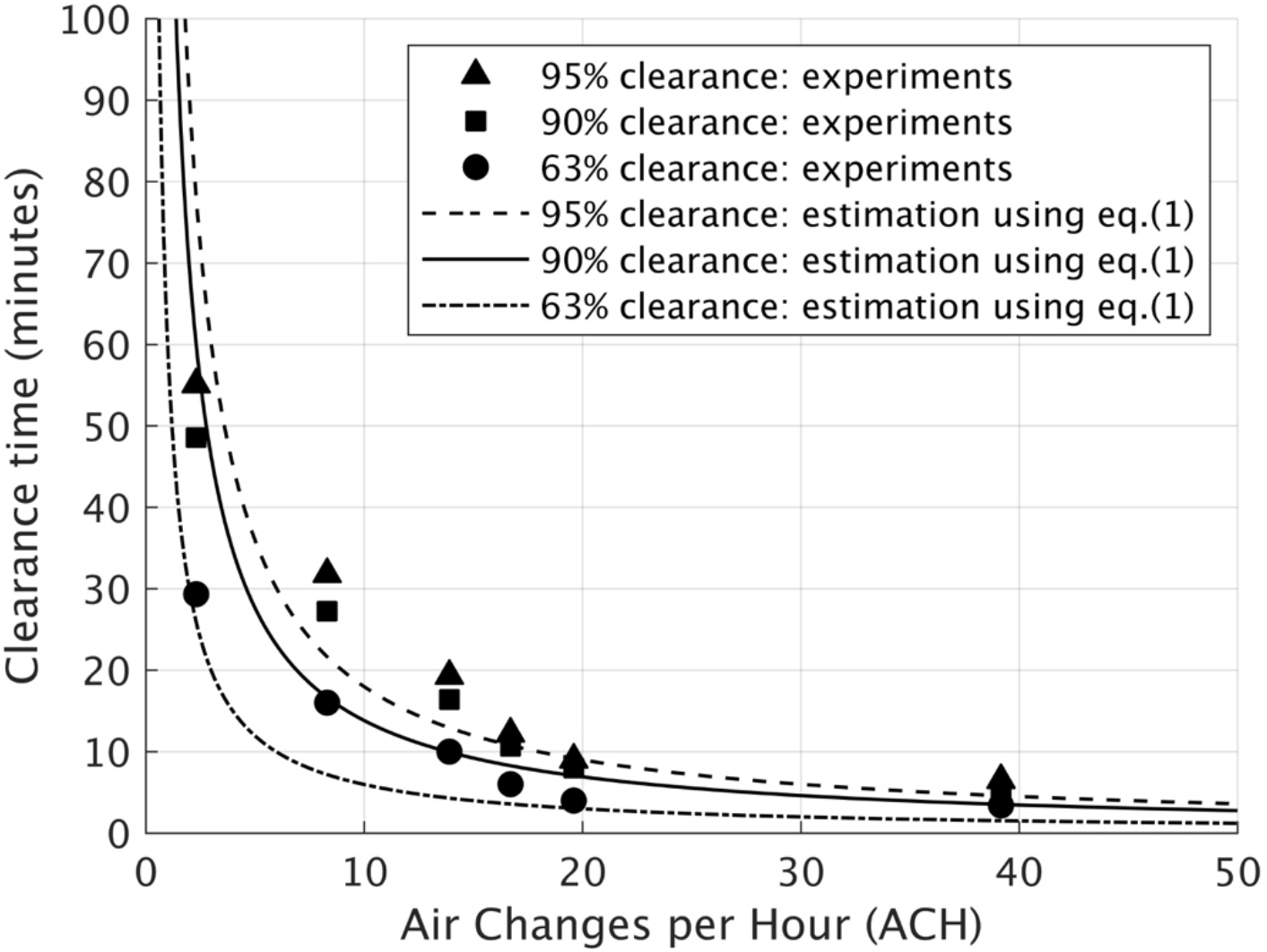
Clearance time as a function of Air Changes per Hour with 63%, 90% and 95% clearance efficiencies. The symbols represent the experimental data for all the cases listed in table 1. The dashed, solid and dot-dashed lines show 63%, 90% and 95% clearance efficiencies, respectively.

## Discussion

In this study, portable air cleaners were shown to be very effective in removing aerosols rapidly by providing high aerosol filtration rates in indoor spaces. The aerosol clearance time results shown in figure 3 indicated that providing sufficiently high ACH for aerosol filtration (∼ 25 ACH), which is not difficult to achieve with portable air cleaners, would reduce the aerosol clearance time significantly. The comparison of the clearance time between the estimations (computed using equation (1)) and the experimental results showed some discrepancy in the low ACH cases (ACH < 15), however, the measured clearance times agreed reasonably well with the estimation when ACH is sufficiently high, such as cases 4 and 6 with 19.6 ACH and 39.2 ACH, respectively. The discrepancy observed in the low ACH cases, such as cases 2 (8.3 ACH) and 5 (13.9 ACH), could be due to the large flow recirculation regions - or “deadzones” - set up in the room that traps the aerosols, delaying the particle motion during the clearance. These recirculation regions likely do not exist at extremely low ACH (e.g. case 1 with 2.3 ACH) because the flow will be entirely laminar and, in any case, the logarithmic decay estimation may not hold at such a low flow rate. This could be because the slow timescale processes such as condensation, leakage through small gaps and weak external pressure variations become relevant during such a slow clearance process. At high ACH, there is likely a sufficient flow rate for homogeneous mixing in a room meaning no significant recirculation regions to delay the clearance of the aerosols. When ACH is relatively low (ACH < 15), there is potentially imperfect mixing due to recirculation regions or other air flow anomalies and the aerosol mixing in the room improves with increasing ACH. It is important to note that the hospital room HVAC (13.9 ACH) remained on while the two air cleaners (25.3 ACH) were tested which provided the total of 39.2 ACH (case 6) when the “air changes” rate of the HVAC (ACH_HVAC_) and two air cleaners (ACHe) were simply combined. As discussed in the introduction, these two types of air ventilation and air filtration methods are fundamentally different such that air changeovers by the HVAC system and “changeovers” by air cleaning devices are not directly equivalent. The role of the HVAC system is to first bring in fresh/filtered air to a room, then circulate this heated/cooled air around a room before exiting it through, or toward, an exhaust vent, thereby pushing out any gaseous pollutants and/or aerosol. Air cleaning devices draw air from inside a room to filter out aerosol and then release aerosol-free air back into the room, while this has the consequence of also circulating air around the room, air circulation is not what the devices are designed for. In the case of both HVAC and air cleaners operating together, as the HVAC circulates air around the room, the strong local flow fields generated by the air cleaners may capture the aerosols before they have the opportunity to travel out of the room. In some cases, it is possible the air cleaners’ rate of aerosol capture could overwhelm the rate of aerosol pushed out of the room by the HVAC. This would only occur when the air cleaner flow rate is sufficiently high relative to the HVAC. A similar explanation was suggested by Miller *et al*. (1996)^12^ based on the findings from their experiments. Further studies are required to prove this postulation but, if correct, this means that portable air cleaners are particularly beneficial in positively pressurised rooms (made positive by design of the HVAC system) that, without such in-room cleaning, serve to push infectious aerosols outside a room potentially reaching susceptible persons in hallways or nursing stations.

Using equation (1) to obtain the clearance time estimation profiles can serve as useful estimation tools for predicting the clearance time for a high clearance efficiency at high ACH, which accounts for mixing effects in a room with inbuilt building HVAC or portable air cleaning devices. However, caution should be used with such an assumption as this does not account for other air flow anomalies such as air flow leakage via room entrances for room geometries that might significantly differ from the rooms studied here. Further work should include investigating a larger room with an assessment of multiple small air cleaners versus smaller number of higher flow rate air cleaners, alongside a study to investigate the best placement of the devices.

## Conclusion

We conclude that standard rates of HVAC air exchanges alone are unlikely to provide sufficient aerosol clearance rate to control aerosol transmission, but a relatively low-cost portable air cleaning devices can dramatically improve the clearance of aerosol indoors in enclosed spaces. Importantly the HVAC system is designed to circulate air flows and relocate the aerosols from one place to another whereas the air cleaners capture and contain the aerosols within the space where those devices are deployed. To clean a room of aerosols in under 10 minutes would require around 25 air changes per hour, which is difficult with HVAC, but is feasible with air cleaning devices.

## Data Availability

The datasets used in the current study were collected by the researchers at the University of Melbourne

## Appendix: Noise level

Realising that putting additional devices into a room could introduce extra un-wanted noise for patients which will be exposed for long periods of time. Tests were performed to measure the noise levels within the clean room when each device was operating. Table 2 shows noise levels for different devices. All devices produce noise at acceptable levels (<65dBA @ 0.5m). Hence, the recommendation is that portable consumer air cleaning devices are employed in high-risk areas when practical.

**Table 2.**
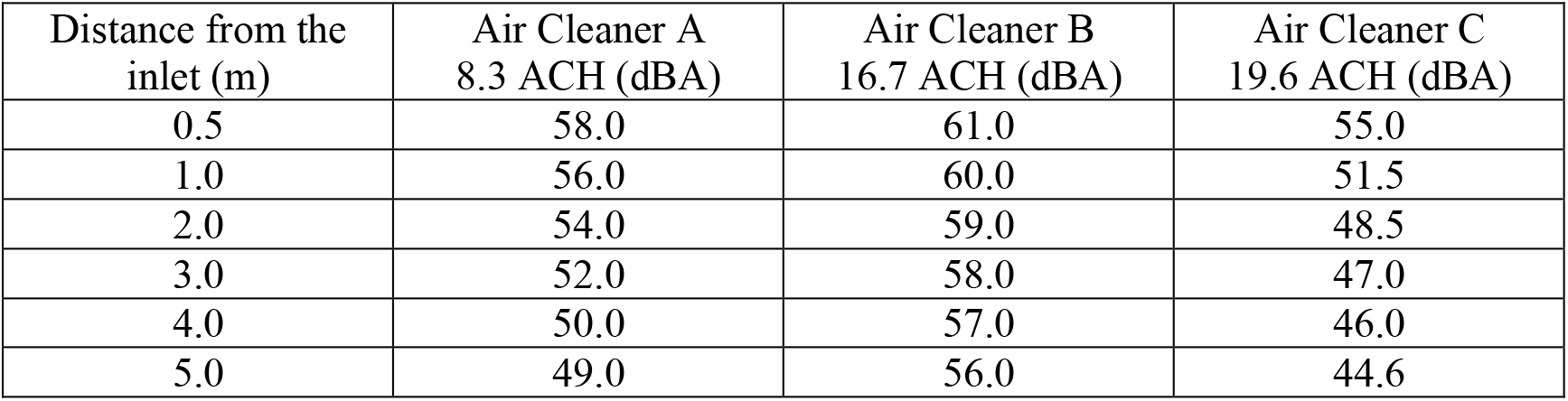
shows noise levels for different devices. Note: Background noise level was 35.5 dBA.

| Distance from the inlet (m) | Air Cleaner A<br>8.3 ACH (dBA) | Air Cleaner B<br>16.7 ACH (dBA) | Air Cleaner C<br>19.6 ACH (dBA) |
| --- | --- | --- | --- |
| 0.5 | 58.0 | 61.0 | 55.0 |
| 1.0 | 56.0 | 60.0 | 51.5 |
| 2.0 | 54.0 | 59.0 | 48.5 |
| 3.0 | 52.0 | 58.0 | 47.0 |
| 4.0 | 50.0 | 57.0 | 46.0 |
| 5.0 | 49.0 | 56.0 | 44.6 |

